# Multimodal Deep Learning Integrating Electrocardiography and Chest Radiography Enhances Prediction of Incident Regurgitant Valvular Heart Disease

**DOI:** 10.64898/2026.07.25.26358904

**Authors:** Siyuan Li, Beijian Zhang, Lei Pan, Zilong Xiao, Ziqing Yu, Juecheng Chen, You Zhou, Zhixing Li, Xiao Li, Cong Wang, Hongyang Lu, Hengli Lai, Yixiu Liang, Junbo Ge

## Abstract

**Background:** Regurgitant valvular heart disease (rVHD) is a major cause of cardiovascular morbidity. Echocardiography is the diagnostic standard but is resource-intensive for large-scale screening. Electrocardiography (ECG) has shown promise for predicting incident rVHD, yet performance varies across phenotypes, particularly for aortic regurgitation (AR). Chest radiography (CXR) provides complementary structural and hemodynamic information. We hypothesized that a multimodal model integrating ECG and CXR would improve prediction of incident moderate-to-severe rVHD.

**Methods:** In this retrospective multicenter study, we identified 212,888 paired ECG-CXR examinations from 116,380 patients across two Chinese centers. Baseline ECG and CXR were obtained within 60 days of echocardiography. Outcome was progression to moderate-to-severe AR, mitral regurgitation (MR), or tricuspid regurgitation (TR). We developed a multimodal neural network with pretrained unimodal encoders, token-level cross-modal fusion, and a class-specific gating mechanism that adaptively weighted ECG-only, CXR-only, and fused predictions. Performance was assessed using C-index, AUROC, AUPRC, decision curve analysis, net reclassification improvement (NRI), and Kaplan-Meier stratification.

**Results:** Multimodal fusion consistently outperformed unimodal models across all phenotypes. For AR, C-index improved from 0.616 (ECG-only) to 0.713 (multimodal; AUROC 0.729, AUPRC 0.972). For MR, multimodal C-index was 0.801 (AUROC 0.814, AUPRC 0.972), versus 0.782 for ECG and 0.775 for CXR alone. For TR, multimodal and CXR-only models showed similar discrimination (C-index 0.802), but multimodal fusion yielded greater net benefit on decision curve analysis. NRI was positive across all time horizons (1-5 years) for all valve types. Grad-CAM interpretability analyses revealed that ECG attention localized to leads II, V₄-V₆ (AR), leads I, II, aVF, V₄-V₆ (MR), and inferior/right precordial leads (TR); CXR attention highlighted chamber-specific enlargement and pulmonary congestion patterns consistent with pathophysiology.

**Conclusion:** A multimodal deep learning model integrating ECG and CXR significantly improved prediction of incident rVHD compared with ECG alone, with the greatest benefit observed for AR. The model leveraged complementary electrical and structural information, demonstrated biological plausibility through interpretability analyses, and provided consistent clinical utility. Given the widespread availability and low cost of both modalities, this approach offers a scalable tool for risk stratification in routine care. Prospective studies are warranted to validate clinical implementation.

## Introduction

Regurgitant valvular heart disease (rVHD) is a major contributor to cardiovascular morbidity and mortality worldwide ^1–3^. Because progressive valvular regurgitation can lead to irreversible chamber remodeling, heart failure, arrhythmia, and adverse long-term outcomes, timely identification of patients at increased risk is clinically important^4,5^. Echocardiography is the reference standard for evaluating valvular structure and severity, but it is resource-intensive and not suitable as a large-scale screening or repeated surveillance tool for all patients in routine practice. More importantly, in rVHD, the clinical value lies not only in recognizing established moderate-to-severe disease, but also in identifying patients at risk before overt progression, when closer follow-up, earlier echocardiographic assessment, and risk management may improve downstream outcomes^6,7^.

Recent advances in artificial intelligence have created new opportunities to extract latent cardiovascular information from widely available routine tests. In particular, deep learning analysis of the electrocardiogram (ECG) has shown promise for detecting structural heart disease and for predicting future valvular dysfunction. Our previous work^8^ has demonstrated that AI-enabled ECG can predict the subsequent development of clinically significant regurgitant valve disease. However, model performance varied across valve phenotypes, with aortic regurgitation (AR) showing weaker discrimination than mitral regurgitation (MR) and tricuspid regurgitation (TR). This limitation is clinically relevant, because it suggests that ECG alone may not fully capture the disease-related signals required for accurate risk stratification in rVHD.

A plausible explanation is that ECG primarily reflects cardiac electrical activity, whereas some of the pathophysiologic changes most relevant to AR are more structural than electrical in nature^9,10^. In this context, chest radiography (CXR) may provide complementary information unavailable from ECG alone, including cardiac silhouette, chamber enlargement, aortic contour, and pulmonary or hemodynamic consequences of valvular dysfunction. From a clinical workflow perspective, ECG and CXR are both inexpensive, widely available, and commonly obtained during the same episode of care, particularly at hospital admission. Their combination therefore represents a practical multimodal approach for scalable risk assessment using tests that are already embedded in routine care.

In this study, we developed and evaluated a multimodal model integrating ECG and CXR to predict future moderate-to-severe AR, MR, and TR. We compared ECG-only, CXR-only, and multimodal models, with discrimination as the primary evaluation domain and decision curve analysis and survival stratification as secondary assessments. We sought to determine whether combining these two routinely available modalities could provide clinically meaningful improvement in risk prediction for rVHD, particularly in disease settings where ECG-based prediction may be intrinsically limited.

## Methods

### Study population

The development cohort was derived from Zhongshan Hospital (Shanghai, China), and included consecutive adult patients aged ≥18 years who received care between July 14, 2017 and December 31, 2023. Eligible patients were identified from routine clinical practice in either inpatient admission or outpatient evaluation settings and were required to have undergone both standard 8-lead ECG, echocardiography and CXR within 60 days, with subsequent echocardiographic follow-up available for outcome ascertainment. This cohort was intended to represent a real-world clinical population in whom ECG and CXR are routinely obtained as part of initial cardiovascular evaluation or general admission workup. The study was approved by the Institutional Research Board of Zhongshan Hospital (B2024-497) with a waiver of patient consent, and registered on ClinicalTrials.gov (NCT07573852).

Overall, the study population was designed as an echo-phenotyped longitudinal cohort anchored to routinely acquired ECG and CXR in real-world care. Baseline echocardiography was used to classify prevalent disease status at enrollment, allowing inclusion of baseline-positive cases during model development while reserving baseline-negative patients for evaluation of future risk prediction.

### Data collection and pre-processing

Raw ECGs were stored as 8-lead signals, including leads I, II, and V1-V6, sampled at 500 Hz for 10 seconds. For each examination, the 8 recorded leads were first reconstructed into a standard 12-lead ECG by deriving leads III, aVR, aVL, and aVF from leads I and II using conventional lead transformation equations. The reconstructed 12-lead ECG was then noise-filtered, harmonic-suppressed and z-score-normalized before model development.

CXRs were originally stored in DICOM format. Each image was also linear intensity corrected and normalized to the range of 0-1 using per-image min-max normalization. Echocardiographic reports were used to define valvular regurgitation status. For each target valve, examinations were classified as positive if the echocardiography report documented moderate or severe regurgitation, and negative otherwise. To construct baseline multimodal samples, all ECGs in the database were first matched against all echocardiograms from the same patient. An ECG was retained if the patient had undergone echocardiography within ±60 days of the ECG, and the valvular status from the temporally closest echocardiogram was assigned as the baseline status of that ECG. For each ECG with an assigned baseline echocardiographic status, all chest radiographs from the same patient were then searched, and a CXR was retained if it had been performed within ±60 days of the ECG. When multiple eligible CXRs were available, only the temporally closest radiograph was selected. Because different ECGs could still be linked to the same CXR, duplicate CXR assignments were further resolved by retaining only the ECG-CXR pair with the smallest absolute time difference. This procedure yielded the final baseline triplets consisting of ECG, CXR, and baseline echocardiographic status.

For each baseline triplet, the ECG acquisition time was defined as the index time. All subsequent echocardiograms from the same patient were then reviewed for follow-up outcome ascertainment. If a positive echocardiographic outcome occurred after the index time, the first positive examination was retained as the event, the follow-up time was defined as the interval from the index time to that event, and the follow-up status was coded as positive. If no positive outcome was observed during follow-up, the last negative echocardiogram was retained as the censoring time, the follow-up time was defined as the interval from the index time to that examination, and the follow-up status was coded as negative, representing right censoring. For samples with a follow-up duration of less than 60 days, the follow-up time was set to 1 day and the follow-up status was considered identical to the baseline status.

The dataset was split at the patient level into a training set and an internal test set at a ratio of 8:2. All examinations from the same patient were assigned to a single dataset to avoid information leakage across partitions.

### Model development

The study outcome was progression to clinically significant rVHD, defined as progression of tricuspid regurgitation (TR), mitral regurgitation (MR), or aortic regurgitation (AR) to moderate or severe during follow-up. We first pretrained unimodal encoders for each modality. For ECG, ECGFounder^11^ was used as the backbone, followed by a prediction head attached to the encoder output to pretrain the ECG branch and generate unimodal ECG predictions. For CXR, Knowledge-enhanced Auto Diagnosis (KAD)^12^ was used as the backbone, with the same strategy applied to pretrain the CXR branch and generate unimodal CXR predictions.

The multimodal model was built on these pretrained encoders and consisted of an ECG branch, a CXR branch, a token-level cross-modal fusion module, and a gated prediction head. The ECG encoder produced a sequence representation of size 20 × 1024 and a 1024-dimensional global feature vector. The CXR encoder produced a token sequence of size 1024*768 and a 768-dimensional global feature vector. To enable cross-modal interaction, ECG and CXR token features were projected into a shared 256-dimensional latent space using linear layers followed by layer normalization and GELU activation. ECG tokens were directly projected from 1024 to 256 dimensions. CXR tokens were first reduced from 1024 to 20 tokens by adaptive average pooling and then projected from 768 to 256 dimensions. Learnable modality embeddings were added to preserve modality identity.

Cross-modal fusion was implemented with two stacked fusion blocks. Each block included modality-specific self-attention, bidirectional cross-attention between ECG and CXR tokens, and modality-specific feed-forward networks with residual connections and layer normalization. Multi-head attention used 8 heads, and the feed-forward expansion ratio was 4.0. After fusion, ECG and CXR tokens were concatenated and pooled using a learnable classification token with multi-head attention. The pooled representation was used as the fused multimodal representation and passed to a multilayer perceptron to generate fused logits. In parallel, unimodal prediction heads were applied to the global ECG and CXR features to generate ECG-only and CXR-only logits.

To combine unimodal and multimodal predictions adaptively, we designed a class-specific gating module. The gating network took as input the original ECG global feature, the original CXR global feature, the fused representation, the projected ECG global feature, and the absolute difference between the projected ECG and CXR global features. For each outcome class, the gating module generated 3 softmax-normalized weights corresponding to the fused, ECG-only, and CXR-only branches. Final logits were computed as the weighted sum of branch-specific logits, followed by sigmoid activation to obtain predicted probabilities. The output layer was formulated in a discrete-time survival framework, allowing the model to learn both time-to-event information and censoring patterns. In this framework, baseline rVHD was encoded at the initial time step, and subsequent time steps represented the probability of incident rVHD during follow-up.

### Model evaluation

Discrimination was primarily assessed using the concordance index (C-index) and the area under the receiver operating characteristic curve (AUROC), and further validated by the area under the precision-recall curve (AUPRC). More intuitively, patients were grouped into low-, intermediate-, and high-risk categories according to model-predicted risk to construct Kaplan-Meier survival curves, comparing event-free survival across the three groups. These metrics were calculated to quantify the ability of the model to distinguish patients who developed clinically significant rVHD from those who did not during follow-up. Clinical utility was assessed using decision curve analysis (DCA), in which the net benefit of the multimodal model was compared across a range of threshold probabilities.

### Model comparison

The performance of the multimodal ECG+CXR model was compared with that of the unimodal ECG-only and CXR-only models using multiple complementary metrics. Discrimination was assessed using the time-dependent C-index and AUROC at the prespecified evaluation horizon (Year 3), allowing direct comparison of predictive performance across modeling approaches. Precision-recall characteristics were evaluated using the AUPRC. In addition, clinical utility was assessed using DCA, in which the net benefit of each model was estimated across a range of threshold probabilities. To further quantify the incremental prognostic value of multimodal fusion beyond ECG alone, both continuous and categorical net reclassification improvement (NRI) were calculated in a time-dependent manner at prespecified horizons (Years 1–5). Continuous NRI was used to assess overall improvement in risk reclassification without predefined categories, whereas categorical NRI evaluated improvement in reassignment across prespecified risk strata. All discrimination, decision-analytic, and reclassification metrics were calculated using the same evaluation dataset and corresponding time horizons for the multimodal and unimodal models. This framework was used to determine whether integration of ECG and CXR provided incremental value beyond either modality alone in prediction of progression to clinically significant rVHD.

### Interpretability analyses

To improve interpretability of the multimodal model, we performed modality-specific and fusion-level analyses. For the ECG branch, two complementary approaches were used. First, gradient-weighted class activation mapping (Grad-CAM) was applied to visualize waveform regions that contributed most to model predictions. Second, a median-beat analysis was performed by ranking patients according to model-predicted risk and selecting the 1,000 highest-risk and 1,000 lowest-risk patients. For each group, median ECG waveforms and corresponding standard deviations were calculated and plotted to summarize characteristic signal patterns associated with extreme predicted risk. For the CXR branch, Grad-CAM was used to visualize image regions that contributed most strongly to model output and to identify anatomically relevant areas associated with risk prediction.

To further examine cross-modal fusion behavior, we performed case studies based on the class-specific gating mechanism of the multimodal model. For selected representative patients and disease-specific models, the branch weights assigned to the fused, ECG-only, and CXR-only prediction pathways were extracted and compared. These analyses were used to illustrate how the relative contribution of each modality varied across patients and across different regurgitant valve phenotypes.

### Statistical analysis

Continuous variables are summarized as mean ± SD, whereas categorical variables are presented as counts with corresponding percentages. Group comparisons for continuous measures were conducted using Student’s t-test, and categorical data were assessed with the chi-square test. For model evaluation, the Likelihood Ratio test was applied to nested Cox models, while the Partial Likelihood Ratio test was used for comparisons between non-nested models. A two-sided p value < 0.05 was considered indicative of statistical significance. All statistical analyses were carried out using Python (version 3.10.12).

### Code Availability

The model weight and code for model training and evaluation are publicly available at Github: https://github.com/zs-xinsight/ECG-CXR-rVHD. Access to the underlying data is restricted due to patient privacy and institutional regulations. Cohort derivation and baseline characteristics

## Results

### Patient characteristics

A total of 212,888 baseline ECG-CXR pairs from 116,380 unique patients were included across all study centers. Baseline characteristics of the study population are summarized in **Table 1**, and cohort derivation is shown in **Figure 1**.

**Figure 1.**
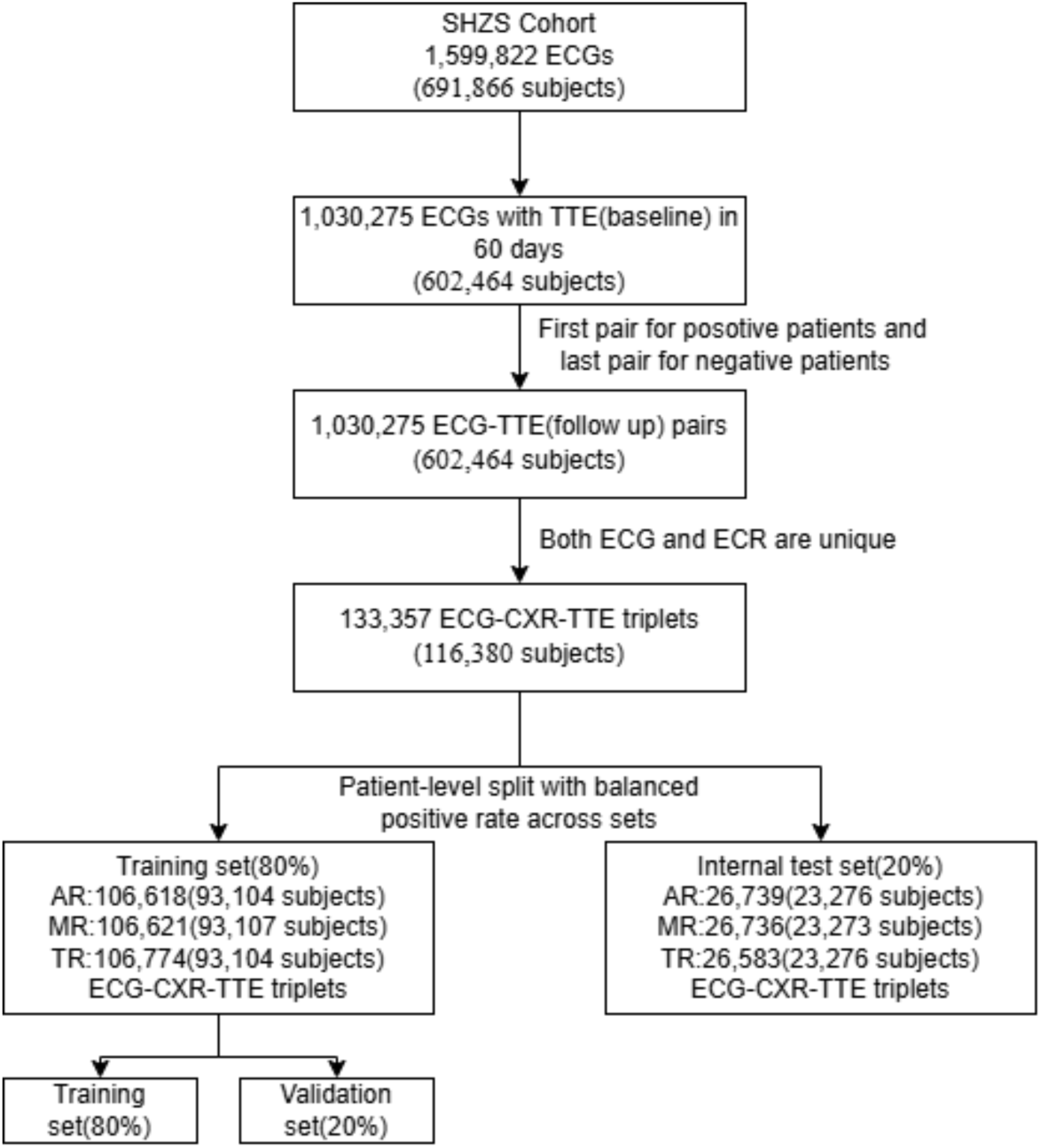
Flow diagram of cohort construction and dataset partitioning. ECG examinations were matched with transthoracic echocardiography and chest radiography to construct unique ECG-CXR-TTE triplets. The cohort was split at the patient level into development and internal test sets, with the development cohort further divided into training and validation subsets. Abbreviations: CXR, chest radiography; ECG, electrocardiography; TTE, transthoracic echocardiography.

**Table 1.**
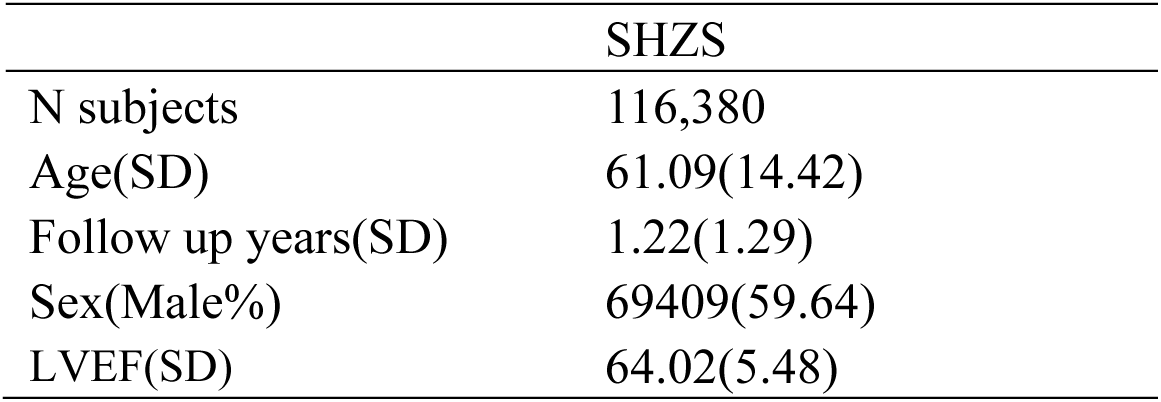
Baseline characteristics of the Zhongshan Hospital study cohort. Continuous variables are presented as mean (standard deviation), and categorical variables are presented as number (percentage).

At Zhongshan hospital, 26,739 paired ECG-CXR samples from 23,276 patients were available for model performance analysis. Among these, 1,658(6.20%), 1943(7.27%) and 1725 (6.47%) pairs were positive at baseline according to the corresponding echocardiographic phenotype, respectively for AR, MR and TR. In patients with no or mild MR, AR, or TR, 5699(22.72%), 5467(22.05%) and 5225(20.94%) pairs had follow-up exceeding 60 days. Among these pairs, 94 (1.64%), 150(2.74%) and 151(2.89%) developed a positive AR, MR and TR outcome during follow-up. The median follow-up duration was respectively 1.36±1.35, 1.28±1.35, 1.34±1.37 years.

Notably, all pairs with positive status at baseline are excluded from the test dataset to present a more realistic predicting ability of our model.

### Primary model performance for prediction of clinically significant rVHD

Model performance varied across valve phenotypes (**Table 2**), with multimodal fusion showing the greatest incremental value for AR. Among the three tasks, the ECG-only model performed worst for AR, with a C-index of 0.616, an AUROC of 0.640, and an AUPRC of 0.957. In contrast, the multimodal ECG+CXR model achieved a C-index of 0.713, an AUROC of 0.729, and an AUPRC of 0.972, outperforming both the ECG-only model and the CXR-only model (0.688, 0.685, and 0.969, respectively). The magnitude of improvement was substantial and statistically significant, indicating that the addition of CXR provided important complementary information for AR risk prediction.

**Table 2.** Discriminative performance of the ECG-only, CXR-only, and multimodal ECG+CXR models for predicting incident moderate-to-severe rVHD. Model performance was evaluated using the concordance index (C-index), area under the receiver operating characteristic curve (AUROC), and area under the precision-recall curve (AUPRC) at the prespecified evaluation horizon. Higher values indicate better performance. Boldface indicates the best-performing model and underlining indicates the second-best-performing model for each valve phenotype and metric. The ECG-only model served as the reference for statistical comparisons. ^#^Reference model; *P<0.05, **P<0.01, ***P<0.001, and ****P<0.0001 versus the ECG-only model.

| Model | C-index↑ | AUROC↑ | AUPRC↑ |
| --- | --- | --- | --- |
| **AR** |  |  |  |
| ECG<sup>#</sup> | 0.616 | 0.64 | 0.957 |
| CXR | <u>0.688<sup>*</sup></u> | <u>0.685</u> | <u>0.969<sup>*</sup></u> |
| ECG+CXR | **0.713<sup>\*\*\*</sup>** | **0.729<sup>\*\*\*</sup>** | **0.972<sup>\*\*\*\*</sup>** |
| **MR** |  |  |  |
| ECG<sup>#</sup> | <u>0.782</u> | <u>0.792</u> | 0.962 |
| CXR | 0.775 | 0.787 | <u>0.965</u> |
| ECG+CXR | **0.801<sup>\*</sup>** | **0.814<sup>\*\*</sup>** | **0.972<sup>\*</sup>** |
| **TR** |  |  |  |
| ECG<sup>#</sup> | 0.752 | 0.739 | 0.956 |
| CXR | <u>0.802<sup>*</sup></u> | **0.796<sup>\*</sup>** | **0.968<sup>\*</sup>** |
| ECG+CXR | **0.802<sup>\*\*\*\*</sup>** | <u>0.793<sup>****</sup></u> | <u>0.962<sup>****</sup></u> |

For MR, the multimodal model also showed the best overall discriminative performance. It achieved a C-index of 0.801, an AUROC of 0.814, and an AUPRC of 0.972, compared with 0.782, 0.792, and 0.962 for the ECG-only model and 0.775, 0.787, and 0.965 for the CXR-only model. Although the absolute gain was smaller than that observed for AR, the multimodal model consistently ranked highest across all discrimination metrics, supporting a modest but reproducible benefit of multimodal integration for MR prediction.

For TR, the point estimates of the multimodal and CXR-only models were similar, but the multimodal model showed greater overall robustness. The ECG+CXR model achieved a C-index of 0.802, an AUROC of 0.793, and an AUPRC of 0.962, compared with 0.752, 0.739, and 0.956 for ECG alone and 0.802, 0.796, and 0.968 for CXR alone. Although the numerical differences between the multimodal and CXR-only models were small, the multimodal model showed stronger statistical significance and more stable performance across evaluation metrics. This pattern suggests that multimodal fusion contributed to more robust risk prediction for TR, even when the improvement in point estimates was modest.

### Clinical utility and risk stratification

Decision curve analysis (**Figure 2**) showed that the multimodal ECG+CXR model provided the highest net benefit across most clinically relevant threshold probabilities for all three valve phenotypes. For AR, the multimodal model began to separate from both unimodal models at approximately the 0.30-0.35 threshold range and maintained the highest net benefit thereafter, remaining above the treat-all strategy once the threshold exceeded roughly 0.35. For MR, differences between models were smaller at low thresholds, but the multimodal model remained consistently higher than ECG alone and slightly higher than CXR alone across much of the 0.20-0.90 range, with clearer advantage over the treat-all strategy from approximately 0.45 onward. For TR, the multimodal model showed the most favorable net benefit over a broad intermediate-to-high threshold range, particularly from about 0.40 to 0.95, where it remained above both unimodal models and exceeded the treat-all strategy after approximately 0.55-0.60. Overall, these findings indicate that multimodal fusion improved threshold-based clinical utility, with the clearest gain observed for AR and TR.

**Figure 2.**
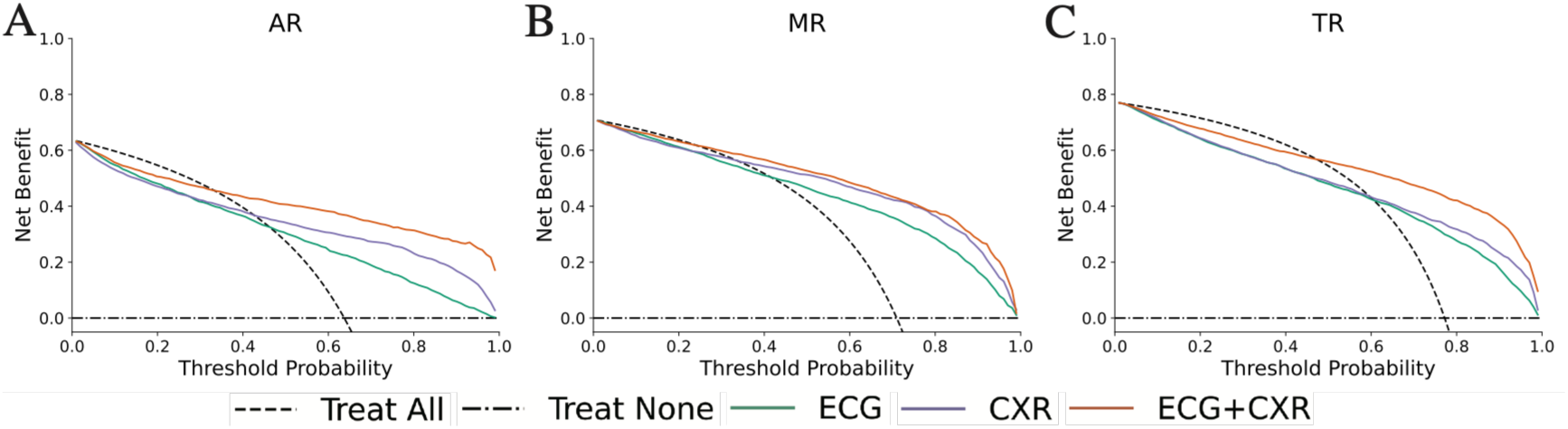
Decision curve analysis of unimodal and multimodal models for predicting incident rVHD. Decision curves compare the clinical net benefit of the ECG-only, CXR-only, and multimodal ECG+CXR models across a range of threshold probabilities for (A) aortic regurgitation, (B) mitral regurgitation, and (C) tricuspid regurgitation. The dashed and dash-dotted lines represent the treat-all and treat-none strategies, respectively.

Kaplan-Meier analysis (**Figure 3**) showed that model-derived risk groups were able to stratify patients into distinct prognostic categories. In the survival plots, panels A, B, and C correspond to the ECG-only, CXR-only, and ECG+CXR models, respectively. Across AR, MR, and TR, patients classified as low risk had the highest event-free survival, those classified as high risk had the lowest event-free survival, and the intermediate-risk group consistently fell between these two extremes. This graded separation was present for all three models, but appeared more distinct and more consistently ordered in the multimodal model, particularly for MR and TR, where the three risk strata remained well separated throughout follow-up. For AR, the multimodal model also showed clearer separation between low-, intermediate-, and high-risk groups than the ECG-only model, indicating improved prognostic stratification when radiographic information was incorporated.

**Figure 3.**
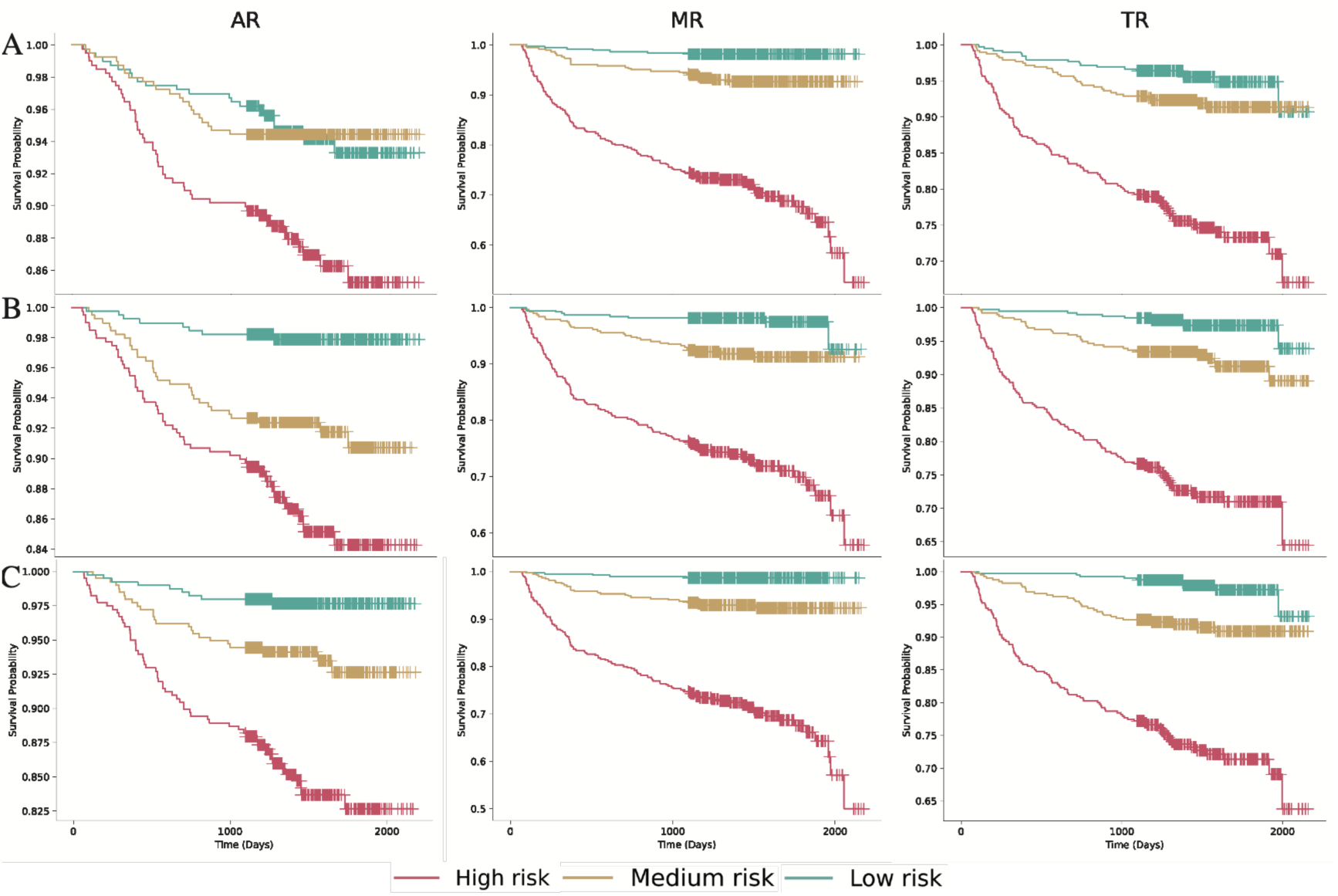
Kaplan-Meier event-free survival curves according to model-defined risk strata. Kaplan-Meier curves show freedom from progression to moderate-to-severe aortic, mitral, or tricuspid regurgitation among patients classified into low-, intermediate-, and high-risk groups. Survival differences among risk groups were assessed by the log-rank test (p<0.0001 for all categories).

Further supporting the clinical utility of multimodal fusion, NRI analysis (**Figure 4**) showed that the ECG+CXR model consistently improved longitudinal risk reclassification compared with the ECG-only model across all valve phenotypes and all evaluated time horizons. Continuous NRI remained positive from Year 1 to Year 5 for AR (0.757–0.594), MR (1.055–0.882), and TR (0.797–0.621), indicating persistent improvement in overall risk assignment over time. Categorical NRI was likewise positive at all horizons, ranging from 0.101 to 0.211 for AR, 0.067 to 0.105 for MR, and 0.118 to 0.134 for TR. Notably, the largest continuous reclassification gain was observed in MR, whereas categorical NRI was particularly favorable in AR and remained stable in TR.

**Figure 4.**
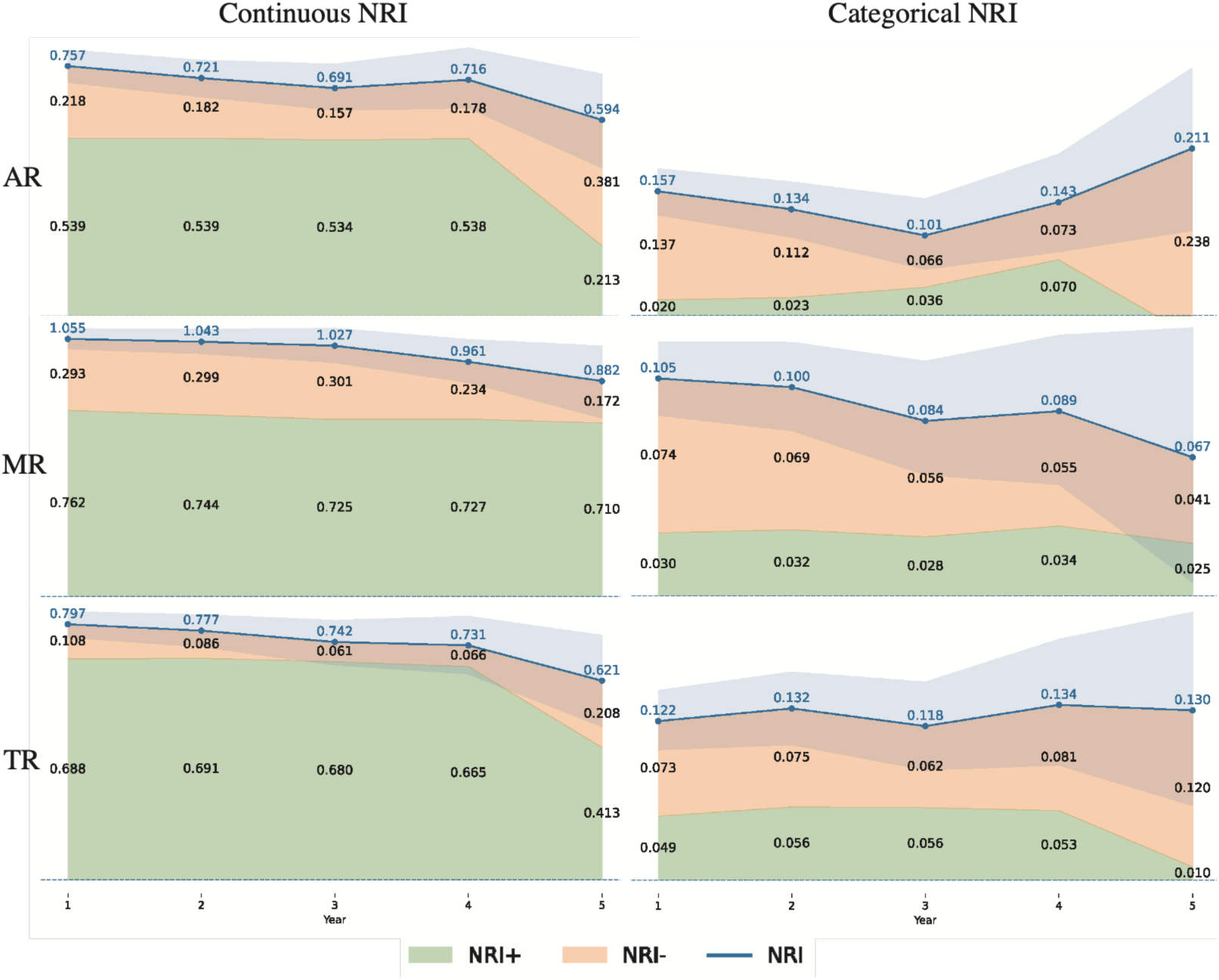
Time-dependent NRI of the multimodal ECG+CXR model compared with the ECG-only model. Continuous and categorical NRI values are shown at 1-, 2-, 3-, 4-, and 5-year prediction horizons for aortic regurgitation, mitral regurgitation, and tricuspid regurgitation. The left column presents continuous NRI and the right column presents categorical NRI based on prespecified risk categories. The blue line represents total NRI, calculated as the sum of NRI+ and NRI-. Positive values favor the multimodal model.

### Subgroup results

Subgroup analyses (**Figure 5**) showed that the multimodal model maintained generally consistent performance across major clinical strata, although the degree of stability differed by valve phenotype. For AR, which was the most challenging task overall, model discrimination remained above 0.60 across all prespecified subgroups, indicating preserved predictive ability even in clinically heterogeneous populations.

**Figure 5.**
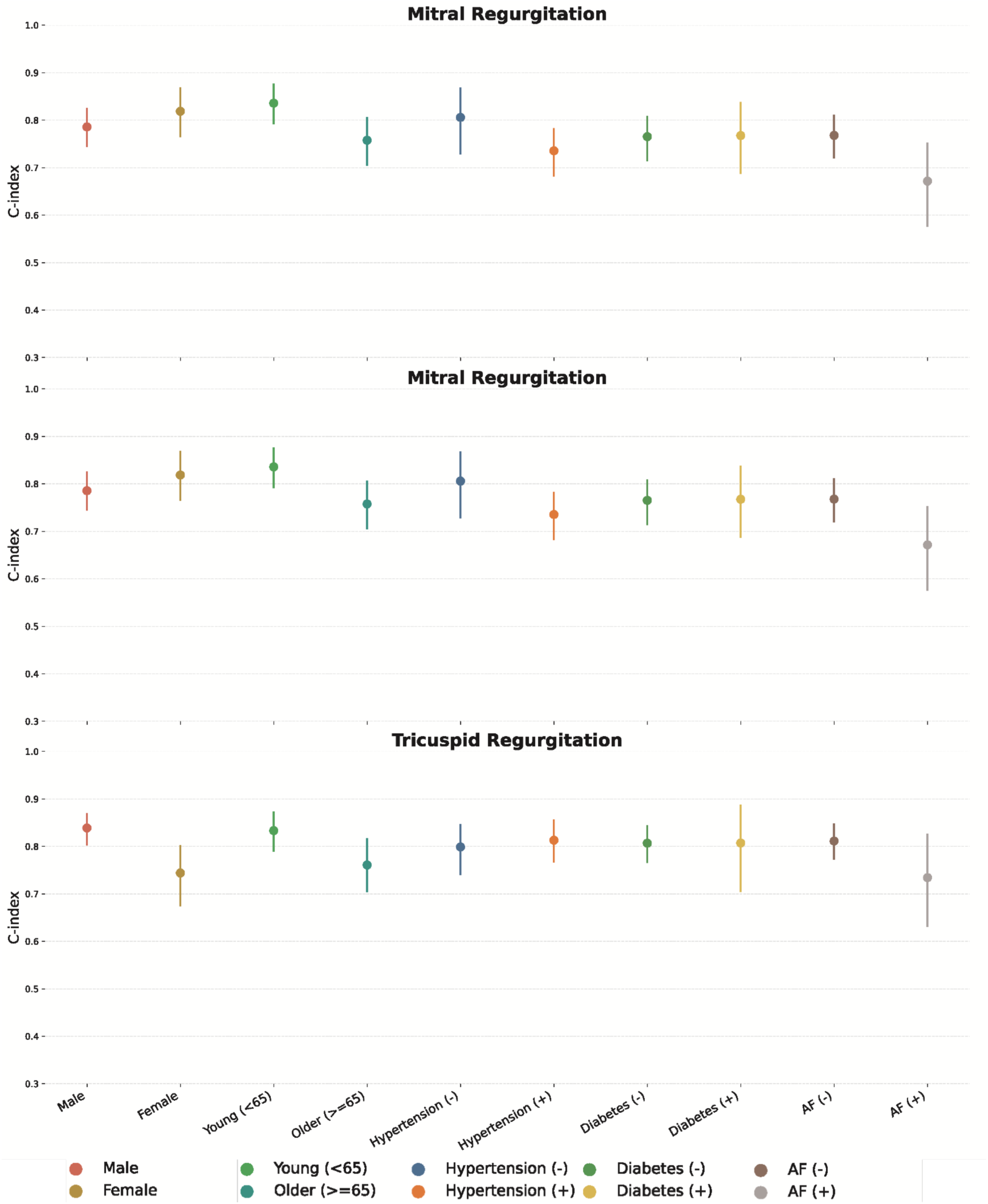
Subgroup performance of the multimodal ECG+CXR model for predicting incident rVHD. C-index point estimates and corresponding 95% confidence intervals are shown for aortic regurgitation, mitral regurgitation, and tricuspid regurgitation across subgroups. Wider confidence intervals in smaller subgroups indicate reduced precision of the corresponding performance estimates.

For MR, model performance was overall more stable, with most subgroup estimates remaining in a relatively narrow and favorable range. Although modest variation was observed across age and comorbidity strata, the general pattern suggested that multimodal prediction for MR was less sensitive to subgroup differences than AR. For TR, subgroup performance appeared both strong and comparatively robust, with less dispersion in point estimates across most strata. Wider confidence intervals were again seen in some comorbidity-positive subgroups, particularly for AF, likely reflecting smaller subgroup size and greater clinical heterogeneity.

### Interpretability analyses

To clarify the contributions of ECG and CXR in predicting incident rVHD, we performed interpretability analyses using gradient-weighted class activation mapping (Grad-CAM) and attention weight quantification for AR (**Figure 6**), MR (**Figure 7**), and TR (**Figure 8**).

**Figure 6.**
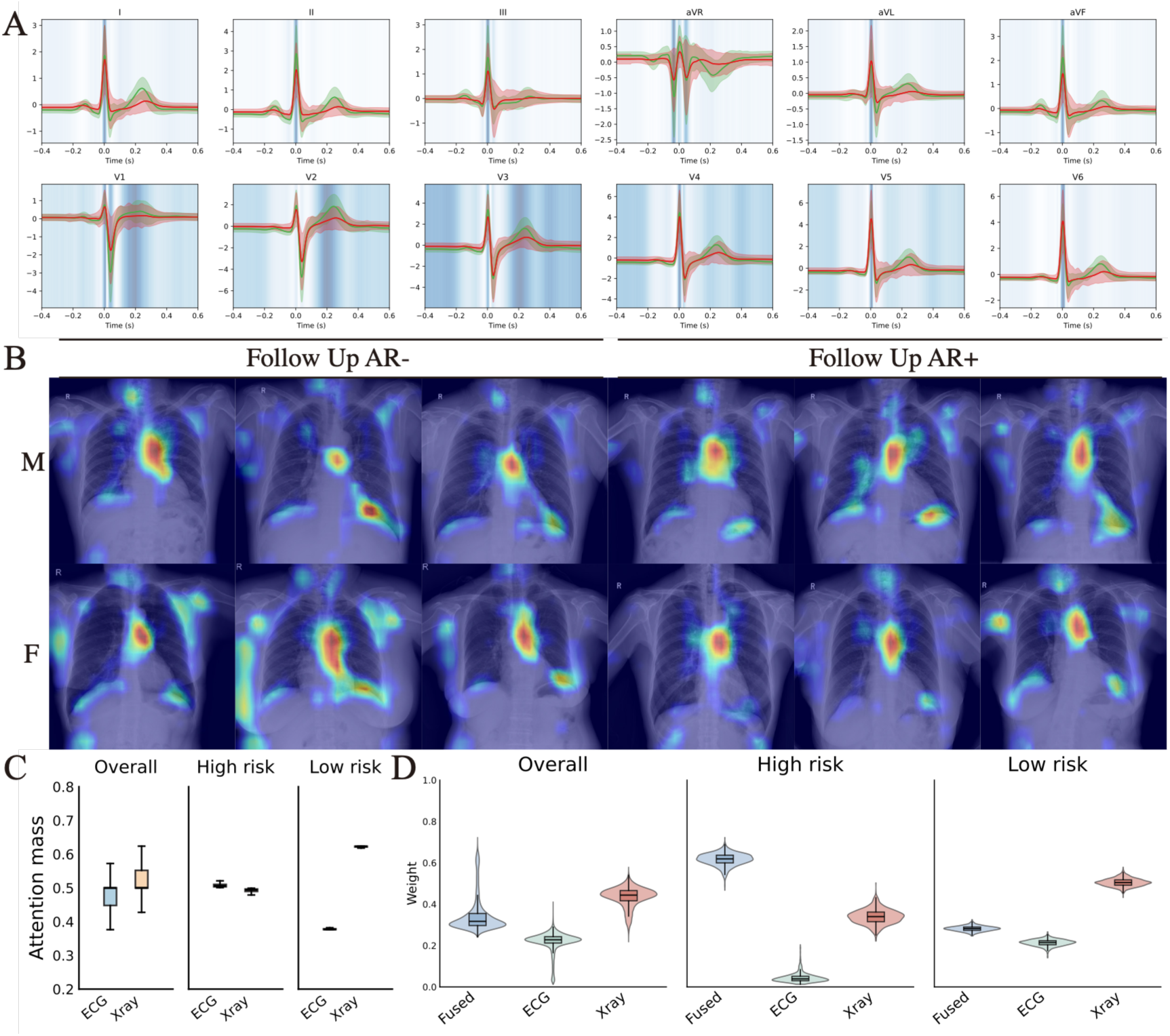
Multimodal interpretability analysis for prediction of incident AR. (A) Median 12-lead ECG waveforms with Grad-CAM attention, highlighting leads II and V4-V6. Shaded bands indicate ±1 standard deviation. (B) Representative CXR Grad-CAM maps stratified by sex and outcome, with increased attention to the left cardiac border and lower lung fields in AR-positive cases. (C) Modality-level attention assigned to ECG and CXR features across the overall, high-risk, and low-risk groups. (D) Gating weights assigned to the fused, ECG-only, and CXR-only prediction branches.

**Figure 7.**
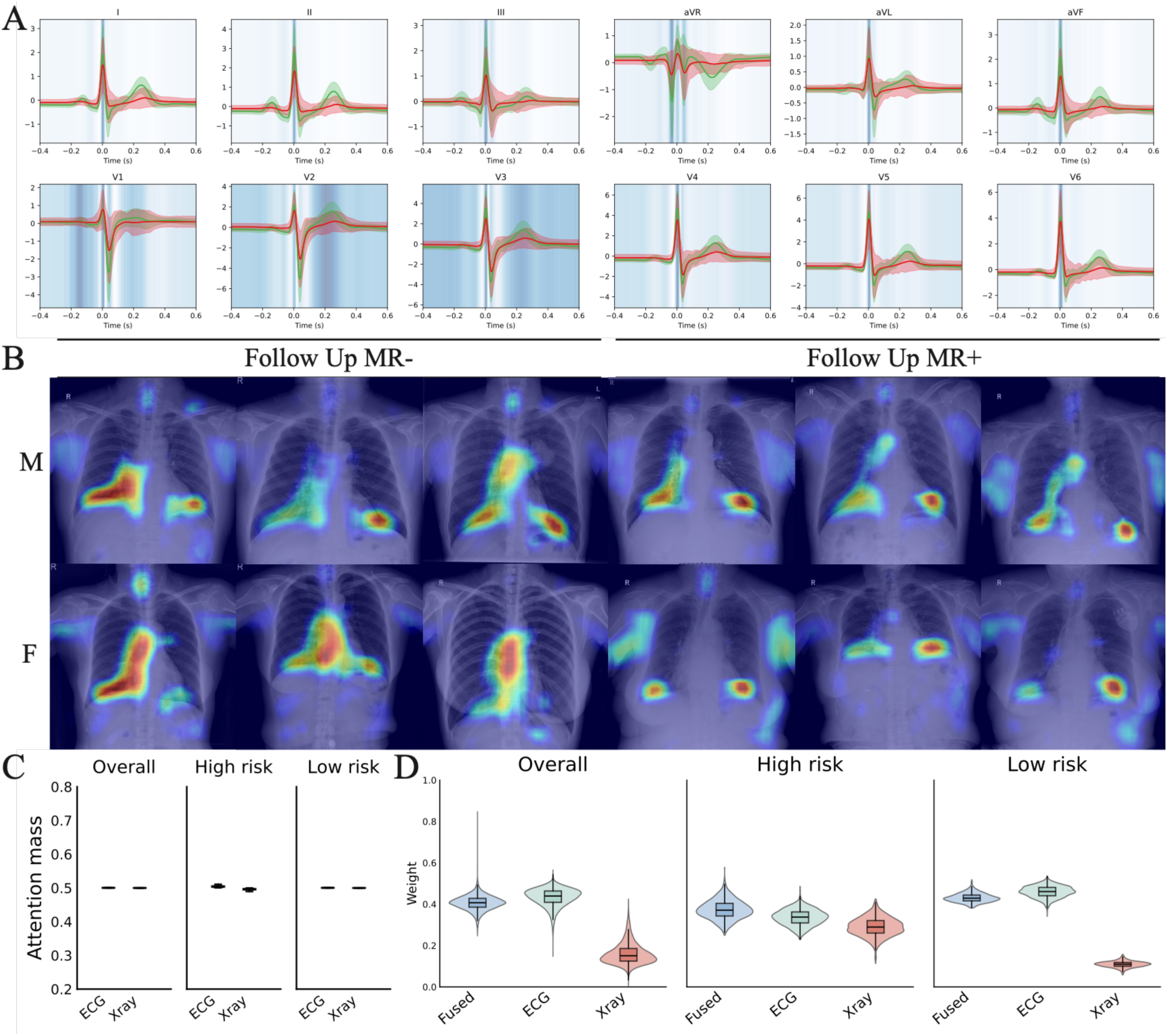
Multimodal interpretability analysis for prediction of incident MR. Panels are arranged as in Figure 6.

**Figure 8.**
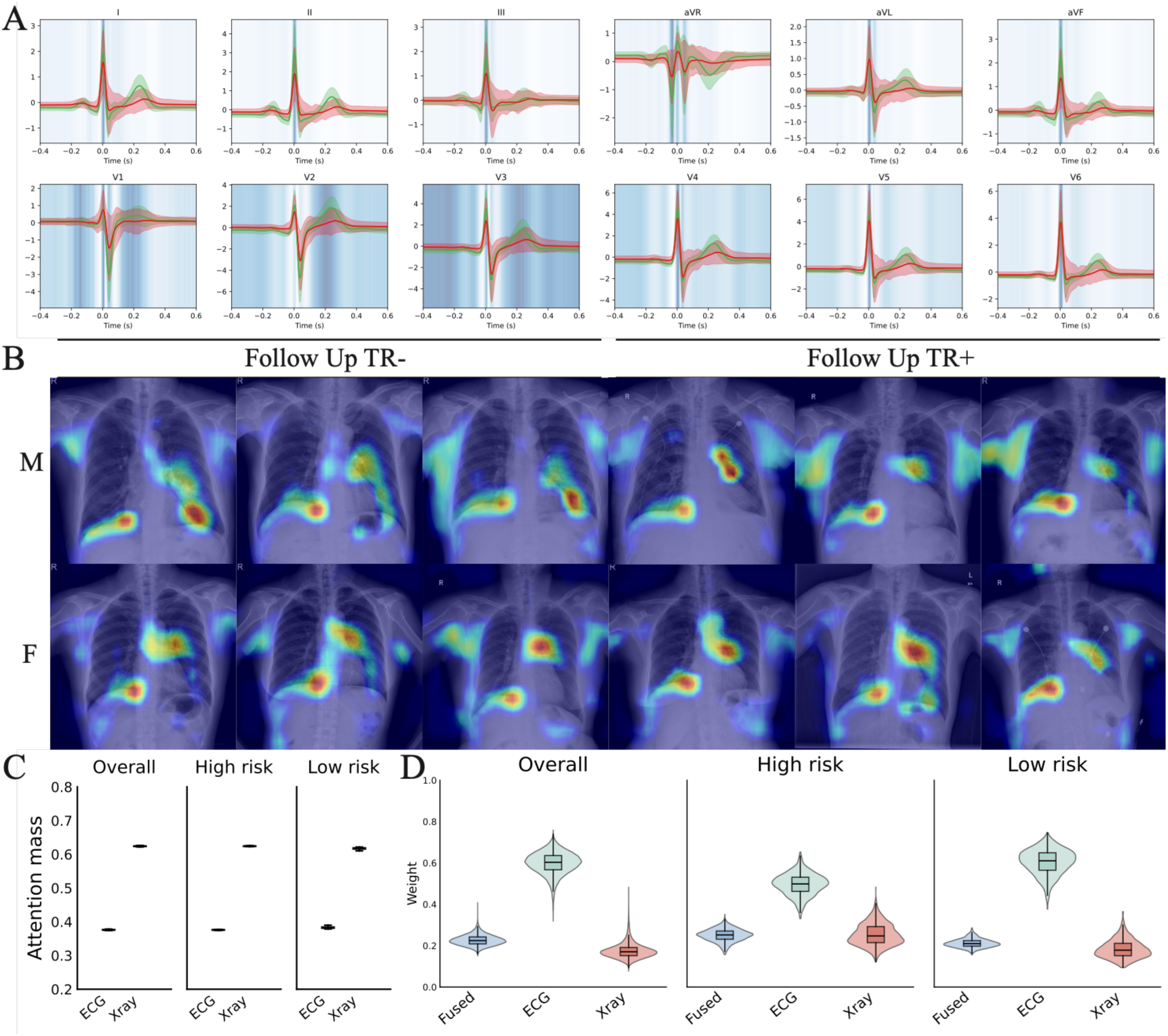
Multimodal interpretability analysis for prediction of incident TR. Panels are arranged as in Figure 6.

ECG Attention Visualization:

For AR (**Figure 6A**), Grad-CAM-generated attention heatmaps of 12-lead ECG revealed distinct activation patterns between patients with incident AR (AR^+^) and those without (AR^−^). Notable differences were observed in leads II, V₄-V₆ (ST-T segment regions), implying that ECG features associated with left ventricular diastolic dysfunction-induced repolarization changes play a role in AR risk stratification. In MR (**Figure 7A**), a pattern of differential activation similar to that in AR was noted in leads I, II, aVF, and V₄-V₆, which may reflect electrical remodeling related to left heart volume overload. For TR (**Figure 8A**), attention was concentrated in the inferior leads (II, III, aVF) and V₁-V₃, consistent with right heart electrical remodeling caused by TR-induced right ventricular dilation.

CXR Attention Heatmaps:

CXR attention heatmaps, stratified by sex (male, M; female, F) and follow-up outcome, provided anatomical insights. In AR (**Figure 6B**), in the AR⁻ group, attention was mainly distributed in the hilar regions, cardiac margins, and mid - lung fields. In contrast, the AR⁺ group showed increased attention in the left cardiac border, pulmonary artery segment, and lower lung fields, which is consistent with left atrial enlargement and pulmonary congestion. Although sex-based differences were subtle, more prominent changes in the left heart silhouette were suggested in females with AR⁺. For MR (**Figure 7B**), in the MR⁻ group, attention was centered on the hilar and basal cardiac regions. In the MR⁺ group, left heart border bulging (indicative of left atrial/ventricular enlargement) and upper lung vascular prominence (suggestive of pulmonary congestion) were observed. Females with MR⁺ had more extensive hyperattenuation in regions related to the left heart. In TR (**Figure 8B**), in the TR⁻ group, attention was in the right hilar and cardiac margin regions. In the TR⁺ group, right cardiac border bulging, pulmonary artery segment prominence, and mid-to-lower lung vascular congestion (consistent with right heart failure) were seen. Females with TR⁺ exhibited more detailed radiological changes related to the right heart.

Quantification of Attention Weights:

In AR (**Figure 6C**), CXR had higher overall attention weights (median ∼ 0.5) than ECG (median ∼ 0.4). However, the weights converged in high - risk subgroups, and the variability of ECG weights increased. In low-risk subgroups, CXR still had a slightly higher weight, but the difference was not significant. For MR (**Figure 7C**), the attention weights of ECG and CXR were balanced overall (median ∼ 0.4–0.5) and across different risk strata, suggesting that they have complementary and stable contributions. In TR (**Figure 8C**), similar balanced weights (median ∼ 0.4–0.5) were observed, with minor shifts favoring CXR in high-risk subgroups or ECG in low - risk subgroups (the differences were not statistically significant). These findings indicate that CXR plays a dominant role in AR (especially in low-risk patients), while ECG and CXR contribute more equitably in MR and TR, and risk stratification modulates their relative importance.

Multimodal Fusion Weights:

The weights of the fusion model (**Figure 6D-8D**) were consistently higher than those of the single-modality models (ECG, CXR) across all three valve diseases and different risk strata, confirming the value of multimodal integration. In AR (**Figure 6D**), the fusion weights (median ∼ 0.9) were the highest overall and in high - risk subgroups. In high-risk subgroups, the weights of ECG exceeded those of CXR, reflecting the enhanced role of ECG in high-risk AR. In MR (**Figure 7D**) and TR (**Figure 8D**), the fusion weights remained the highest, and the weights of ECG and CXR were more balanced (median ∼ 0.3-0.4), which is consistent with their equitable contributions.

## Discussion

We developed the first bimodal model integrating ECG and CXR to predict future progression of moderate-to-severe rVHD, and evaluated its performance across multicenter cohorts from different regions. Importantly, the addition of the chest radiography modality significantly improved the performance of the ECG-only model, likely by capturing complementary structural not fully reflected by electrical signals alone, thus providing biologically plausible prediction of disease progression. Our findings suggest that fusion of ECG and CXR information may be used to guide surveillance echocardiography in patients at risk of future rVHD progression, thereby facilitating earlier prevention and intervention.

### ECG+CXR Bimodal Model Efficiently Predicts rVHD Progression

The multimodal ECG+CXR model consistently outperformed both unimodal models across all three regurgitant phenotypes. For AR, the improvement was substantial: the C-index increased from 0.616 (ECG only) to 0.713 (multimodal), representing a 15.8% relative improvement. For MR and TR, the gains were more modest but still present. Notably, the multimodal model achieved the highest net benefit in decision curve analysis and maintained favorable survival stratification throughout follow-up. NRI was positive at all evaluated time horizons (1–5 years) for all valve types, underscoring the incremental prognostic value of adding CXR to ECG-based prediction. These results remained consistent in both internal and external validation cohorts, supporting the robustness and generalizability of the integrated approach. The ability of the model to derive risk estimates from routine, non-invasive tests that are already available during a single episode of care positions it as a practical tool for large-scale risk stratification and clinical workflow integration^13^.

### Biological Rationale for ECG- and CXR-Based AI Prediction of rVHD Progression

Progressive valvular regurgitation induces both electrical and structural cardiac remodeling before overt clinical symptoms emerge. ECG reflects electrical consequences such as left ventricular hypertrophy, left atrial enlargement, and repolarization abnormalities, which have long been associated with valvular disease ^14,15^. In AR, volume overload leads to left ventricular dilation and diastolic dysfunction, often manifesting as ST-T changes in lateral leads. In MR, left atrial volume overload produces P-wave prolongation and atrial fibrillation, both detectable on surface ECG^16^. In TR, right ventricular pressure overload can give rise to right axis deviation and right-sided chamber enlargement patterns on ECG.

Chest radiography provides direct anatomic and hemodynamic information: cardiac silhouette enlargement, chamber-specific dilation (e.g., left atrial double right border in MR, right atrial bulge in TR), pulmonary vascular redistribution, and signs of congestive heart failure^17,18^. These morphologic signs are often present even when ECG remains normal or non-diagnostic, particularly in early-stage AR. The biological complementarity between the two modalities, electrical versus structural, provides a strong rationale for their combined use in AI-based risk prediction of rVHD.

### Interpretability Analyses Support the Biological Plausibility of the ECG-CXR Model

Grad-CAM attention maps revealed that the model’s decision-making aligned with known pathophysiologic patterns. For AR, ECG attention concentrated on leads II, V₄–V₆ and the ST-T region, corresponding to left ventricular repolarization abnormalities commonly seen in chronic aortic volume overload. CXR attention in AR-positive patients highlighted the left cardiac border, pulmonary artery segment, and lower lung fields, consistent with left atrial enlargement and pulmonary congestion. For MR, the model consistently attended to leads I, II, aVF, and V₄-V₆ on ECG (reflecting left atrial and left ventricular electrical remodeling) and the left heart border and upper lung vasculature on CXR (indicative of left atrial enlargement and pulmonary venous hypertension). For TR, the ECG focus shifted to inferior leads (II, III, aVF), matching the electrophysiologic signature of right ventricular hypertrophy or dilation; CXR attention was high over the right heart border and pulmonary artery, consistent with right heart chamber enlargement. The sex-stratified heatmaps further showed subtle differences, with female patients exhibiting more pronounced changes in left heart structures in AR and MR, in agreement with known sex disparities in valvular disease presentation^19^. These interpretability analyses provide face-valid evidence that the model leverages clinically recognizable features rather than spurious correlations.

### Inclusion of the CXR Modality Yields Varying Degrees of Performance Improvement across Different rVHD

The incremental value of CXR differed strikingly across valve phenotypes. The greatest relative improvement was observed in AR, where the ECG-only model performed worst (C-index 0.616) and the addition of CXR raised discrimination to 0.713. This large gain underscores the fundamental limitation of ECG alone in detecting early AR: the electrocardiogram often remains normal until advanced stages, whereas chest radiographs can demonstrate aortic root dilation, left ventricular prominence, and pulmonary congestion even when ECG findings are equivocal^10,20,21^. In MR, ECG alone already achieved a C-index of 0.782, and the multimodal model provided a statistically significant but numerically modest improvement (to 0.801). The strong performance of ECG in MR likely stems from the high frequency of left atrial electrical abnormalities (P-wave abnormalities, atrial fibrillation) that are readily captured by surface leads; CXR added complementary structural information (left atrial enlargement, pulmonary venous redistribution) that further refined risk stratification. For TR, the CXR-only model performed equally well as the multimodal model (both C-index 0.802), and the multimodal addition did not improve point estimates. This may be because TR produces prominent right heart structural changes (right atrial/ventricular enlargement) that are easily detected on CXR, and the electrical correlates of TR on ECG are less sensitive; thus, the CXR already carries most of the prognostic signal. Nevertheless, the multimodal model demonstrated greater net benefit on decision curve analysis for TR, suggesting that its main added value lies in improving clinical utility, particularly in reducing the number of false-positive alerts, rather than in further advancing conventional discrimination metrics.

The adaptive nature of the multimodal model, which employs a class-specific gating module that dynamically weighs ECG, CXR, and fused representations, allowed it to capture the optimal complementary emphasis for each valve type. For AR, the model gave higher weight to CXR features in low-risk patients and enhanced ECG contributions in high-risk patients, as shown by the attention weight analysis (**Figures 6C-D)**. For MR and TR, the weights were more balanced, reflecting the already strong performance of each single modality. This ability to adaptively weight modalities based on risk subgroups and disease phenotype is a key strength of our gated fusion design, enabling the model to leverage the “best of both” depending on the clinical context.

### ECG and CXR Data Are Highly Accessible in Clinical Practice

ECG and CXR are particularly attractive modalities for multimodal model development and clinical deployment because, as shown in our study, they are routinely obtained early during hospitalization as part of initial cardiovascular evaluation or general admission workup, and are commonly available within the same episode of care. This high degree of accessibility enabled the assembly of a large real-world paired ECG–CXR dataset for robust model training and validation. More importantly, these two modalities provide complementary rather than redundant information: ECG primarily captures electrical manifestations of myocardial and conduction abnormalities, whereas CXR provides structural and hemodynamic information (cardiac silhouette, chamber enlargement, aortic and pulmonary changes) that cannot be obtained from electrical signals alone. From an implementation perspective, ECG is the most commonly ordered cardiac test, and chest radiography is among the most frequently performed imaging examinations^22,23^, making this inherently scalable with minimal marginal cost. In practice, such a workflow could allow future risk of rVHD progression to be estimated directly from routinely acquired admission data, thereby supporting in-hospital risk stratification, prioritization of follow-up echocardiography, and broader patient management. This type of workflow integration is clinically plausible, as multisite randomized trials have already shown that AI-enabled ECG alerts embedded in routine hospital care can identify high-risk patients and reduce all-cause mortality^24,25^.

### Clinical Implications

The ability to predict incident moderate-to-severe rVHD using only baseline ECG and CXR, two low-cost, widely available tests, has substantial clinical implications. Current guidelines recommend echocardiography for patients with signs or symptoms of valvular heart disease, but many individuals at high risk remain undetected until advanced stages, when irreversible myocardial damage has already occurred. Our model could be integrated into the electronic health record as a real-time screening tool: when a patient undergoes ECG and CXR for other reasons (such as admission for pneumonia, surgical evaluation, or routine annual check-up), the model could calculate a risk score for each regurgitant phenotype. Patients identified as high-risk could be flagged for earlier echocardiography and closer cardiology follow-up. This approach would be particularly valuable for AR, which remains the most challenging to detect using conventional tools. Furthermore, the decision curve analysis demonstrates that the multimodal model provides net benefit across a wide range of clinically relevant thresholds, suggesting that it could help avoid unnecessary echocardiograms in low-risk patients while capturing high-risk patients who are currently missed. Prospective studies are warranted to evaluate whether incorporating this AI-guided strategy into clinical workflows improves downstream cardiovascular outcomes.

### Limitations

Several limitations should be acknowledged. First, this was a retrospective study based on routine clinical data and is therefore subject to inherent selection bias and residual confounding. Because inclusion required the availability of ECG, CXR, and follow-up echocardiography, the study population may not fully represent the broader population of patients with rVHD encountered in routine practice. Second, although the model was evaluated across multicenter cohorts, all data were derived from domestic centers, without validation in geographically or ethnically distinct populations. This may limit the generalizability of the present findings to other healthcare systems, imaging workflows, and population structures. Third, outcome ascertainment relied on echocardiographic reports interpreted by clinicians rather than centralized core-laboratory adjudication. Accordingly, some degree of inter-observer variability and misclassification is unavoidable, particularly for borderline cases or subtle interval progression. Fourth, the ECG input was reconstructed from 8 recorded leads to standard 12-lead format; although lead transformation is a well-validated technique, some information loss may occur. Fifth, we only evaluated incident rVHD and did not examine progression of pre-existing disease or valvular stenosis. Sixth, the event rate was low (1–3% of eligible samples), leading to class imbalance that may inflate metrics such as AUPRC. Finally, while the present model showed encouraging performance in discrimination, risk stratification, and clinical utility analyses, its real-world value in guiding surveillance echocardiography and improving patient outcomes remains to be established. Prospective studies, ideally in the form of a pragmatic clinical trial, are needed before this approach can be considered for clinical implementation.

## Conclusion

In this multicenter study, we developed and validated a multimodal deep learning model integrating electrocardiography and chest radiography to predict future progression of aortic, mitral, and tricuspid regurgitation to moderate-to-severe severity. The integration of chest radiography provided complementary information that significantly enhanced the predictive performance of ECG alone, especially for aortic regurgitation. Interpretability analyses confirmed that the model’s attention aligned with disease-specific electrical and structural aberrations. The dynamic gating fusion mechanism allowed the model to adaptively balance the contributions of the two modalities according to disease phenotype and risk subgroup. Given the worldwide availability and low cost of ECG and CXR, this multimodal approach offers a practical and scalable tool for the early identification of patients at high risk for clinically significant regurgitant valvular heart disease, potentially enabling earlier echocardiographic surveillance and intervention. Further prospective studies are warranted to assess its clinical utility and impact on patient outcomes.

## Clinical Trial Number

ClinicalTrials.gov (NCT07573852)

## Data Availability

Access to the underlying data is restricted due to patient privacy and institutional regulations. Cohort derivation and baseline characteristics.

## Notes

### Competing Interest Statement

The authors have declared no competing interest.

### Author Declarations

Ethics committee/IRB of Zhongshan Hospital gave ethical approval for this work

### Funding

- Shanghai Pujiang Talent Program: 24PJD014
- AI for Science Foundation of Fudan University: FudanX24A1056

